# Risk factors for enteric pathogen detection in food consumed by children aged 6–24 months in informal urban neighbourhoods of Nairobi, Kenya: a cross-sectional study

**DOI:** 10.1101/2025.08.28.25334656

**Authors:** Lynn Grignard, Jackie Knee, Alesha Myers, Joseph Wells, Julia Sobolik, Clara MacLeod, Linnet Ochien’g, Christine Mutisya, Sherril Masudi, Joseph Wasonga, Alice Kiarie, Noah Okumu, Delia Grace, Johanna Lindahl, Maud Amon-Tanoh, Julie Watson, Elizabeth A. J. Cook, Oliver Cumming

## Abstract

**Background:** Food is an important transmission route for symptomatic and asymptomatic enteric infections in early childhood. Chronic carriage of enteric pathogens may lastingly impact gut health and childhood growth and development. This cross-sectional study aims to describe the detection frequencies of 30 enteric pathogens in food samples using TaqMan Array Cards and their associated risk factors among children aged 6–24 months living in a low-income, densely populated urban area of Nairobi, Kenya.

**Methods:** We conducted a cross-sectional study among 569 households in the Dagoretti Division of Nairobi, Kenya. We collected one food sample from each household and tested each sample for 30 enteric pathogens by custom TaqMan Array Card. The relationship between potential food-related risk factors and the detection of enteric pathogens in food was explored using multivariable logistic regression.

**Results:** The prevalence of detection of ≥1 enteric pathogen found in children’s food was 21.4% (n=122/569). The most prevalent pathogens were *Aeromonas* (9%, n=51/569) and *Cryptosporidium* (5.3%, n=30/569). Caregivers selecting food vendors based on hygiene was associated with decreased odds of detecting ≥1 protozoan pathogen (OR 0.55, 95% CI 0.30, 0.99, p=0.049). Preparing food in a container was found to be associated with decreased odds of bacterial detection (OR 0.56, 95% CI 0.33, 0.96, p=0.033), largely driven by detecting *Aeromonas* (OR 0.37, 95% CI 0.19, 0.71, p=0.003). Rodent sightings in the household within the last week were associated with increased odds of detecting *Cryptosporidium* in child food (adjusted odds ratio [OR] 2.64, 95% confidence interval [CI] 1.10, 7.37, p=0.042). Conversely, rodent sightings were associated with a reduced risk of detecting bacteria in food (adjusted odds ratio [OR] 0.60, 95% confidence interval [CI] 0.36, 0.99, p=0.044.

Mixtures of cereal, meat, fish, vegetables, and legumes were weakly associated with increased odds of detecting bacteria (OR 1.69, 95% CI 0.97, 2.94, p=0.063) in child food, mostly driven by the detection of *Aeromonas* (OR 2.25, 95% CI 1.18, 4.35, p=0.015). Food containing dairy was associated with decreased odds of detecting ≥1 enteric pathogen in the sample (OR 0.42, 95% CI 0.21, 0.78, p=0.009) even though the numbers were small.

**Conclusion:** We detected a range of enteric pathogens in food consumed by children aged 6-24 months of age in this setting. Our results suggest that contaminated food may be an important pathway of disease transmission among young children. Targeted food hygiene interventions are needed to address these specific foodborne risks for young children in complex urban settings such as this.

## Introduction

Contaminated food is an important route of transmission for enteric pathogens (1). Globally, there are an estimated 600 million cases of foodborne illnesses each year, with children under five bearing 40% of this burden. The burden is highest in low-and middle-income countries (LMICs), where food contamination levels are frequently high (1–3). The East African region, where Kenya is situated, is reported to have the second-highest level of foodborne disease globally (4).

Two-thirds of all diarrhoeal deaths occur in children under two, with over 50% occurring in children between 6–11 months of age (5). This coincides with the weaning period when children become exposed to complementary foods, protection from maternal antibodies declines and immunity from breastmilk wanes, leaving children under two particularly vulnerable to infection (2, 6–8). Infections with enteric pathogens often present with gastrointestinal symptoms such as diarrhoea, nausea, and vomiting. Repeated episodes of diarrhoea reduce nutrient absorption and increase nutrient loss, leading to malnutrition, a weakened immune system, and a higher risk of infection (9). Chronic carriage of both symptomatic and subclinical enteric infections may lastingly impact gut health and childhood growth and development (10, 11).

Contamination of food can occur at various points along the farm-to-fork continuum, from agricultural production and processing to domestic food hygiene practices (5). In high-income countries (HICs), foodborne transmission of enteric pathogens is closely regulated by authorities to ensure safety throughout the food supply chain. In contrast, food safety monitoring in LMICs is often insufficient. Unsanitary conditions and lack of regulation at farms and markets can lead to contamination of milk and other food types with human or animal faeces before household use (1, 12, 13). Additionally, poor domestic hygiene practices such as the use of unsafe water for preparing and washing foods, insufficient handwashing, poor sanitation in food preparation areas, failure to cook infant foods to safe temperatures, improper storage of perishable items at ambient temperatures, and exposure to flies due to uncovered food containers can all contribute to increased microbial contamination (14–18). Recent interventions are now focusing on targeted ways to improve domestic child food hygiene by reducing the number of pathogens present and improving the overall microbiological food quality (7, 19).

To date, studies exploring risk factors for food contamination in LMICs have mainly used traditional culture-based methods. These conventional methods for foodborne pathogen detection use *Escherichia coli* and other faecal indicator organisms and measure the colony-forming units per g of food (CFU/g), as indicators of microbial contamination of child food (2). Nucleic acid-based methods, such as quantitative multiplex PCR, isothermal amplifications, and biosensor-based methods, sensitively and rapidly detect multiple foodborne pathogens (9, 20, 21). They are now widely used in the food safety industry in high-income countries to prevent foodborne disease outbreaks (22). Advanced molecular methods should be implemented in LMIC as they are more sensitive than traditional methods, important foodborne viruses do not correlate with the presence of *E. coli*, and the field is moving towards understanding specific pathogen impact on health rather than correlates of contamination.

Foodborne diseases, caused by the ingestion of food or beverages contaminated with pathogenic microorganisms, represent a significant public health threat in young children in LMICs. It is essential to rigorously evaluate domestic child food hygiene practices to identify critical areas for targeted interventions. This cross-sectional study describes the detection frequencies of 30 enteric pathogens in food samples using TaqMan Array Cards and their associated risk factors among children between 6–24 months living in a low-income, densely populated urban area of Nairobi, Kenya.

## Methods

### Study design and setting

We conducted a cross-sectional study in Dagoretti South Sub-County, Nairobi, Kenya, as part of the multi-country Urban Infant Foodscape (UIF) study (23, 24). We collected and analysed food samples to report the prevalence of enteric pathogens and their associated risk factors in the Dagoretti South Sub-County, Nairobi, Kenya.

Dagoretti South Sub-County is a peri-urban, low-income settlement within Nairobi City County, Kenya. The sub-county covers 29.1km^2^, comprising 9.87% (434,208) of Nairobi County’s population. It is characterised by a high infectious disease burden, inadequate Water, Sanitation and Hygiene (WASH) services, and a high population density (17). Dagoretti South Sub-County is divided into five administrative units or wards. Households were recruited from two wards – Uthiru/Ruthimitu and Riruta – for this study based on their proximity to health facilities.

### Enrolment

Households were eligible to participate in the study if they had at least one child aged between 6–24 months living in the household at the time of the study, the primary caregiver of the child provided written informed consent to both their and the child’s participation. Trained field enumerators conducted all enrolment, data and sample collection activities in English or Kiswahili between June 2021 and September 2021. To recruit households, a sample frame was created using the community health volunteers (CHVs). From the 259 CHVs in the study wards, 90 were randomly selected to participate in the study. The CHVs mapped every household with a child between 5-23 months in the two wards in Dagoretti South Sub-County selected for the study. Households identified to have at least one child aged between 5–23 months (to ensure they were still within the eligible age range at the time of data collection) were included in the sample frame. From this sample frame, households were randomly selected for participation using Microsoft Excel. Where a household refused to participate, was not available during the sampling period (3 consecutive days), or was found to be ineligible, another household from the sample frame was randomly selected.

### Data and food sample collection

Field teams visited each participating household and, after gaining consent, verbally administered a detailed household survey to the primary caregiver of the index child (one child aged between 6–24 months) to collect data on socio-demographic characteristics of the household, access to services and behavioural practices related to water, sanitation, and hygiene (WASH), the child’s dietary habits and recent illness, contact with animals and other general household and child-level risk factors. All data were collected using Open Data Kit on electronic tablets and uploaded onto a dedicated, encrypted server at the end of each data collection day. Research teams at both ILRI and the London School of Hygiene and Tropical Medicine (LSHTM) cross-checked the data daily. For the food sample collection, caregivers were asked to give a sample of food that they would feed to the child if they were hungry. With a plastic tablespoon, two spoonsful of food were collected into 50ml Falcon tubes, labelled, and placed in a rack inside a cool box. Cool boxes were collected twice daily (11:00 am and 2:30 pm) and transported to the laboratory for processing.

### Laboratory analysis

Each food sample was homogenised in a stomacher for 2 minutes and shaken by hand 25 times to ensure an even distribution of microorganisms. 2.5 g aliquots of the homogenised food were stored at-20°C ready for extraction. Total nucleic acid was extracted from food samples using a CTAB (cetyltrimethylammonium bromide) based method (26). Briefly, 200mg of pre-homogenised food, 1mL CTAB lysis buffer (Cat #A4150.0500, 20 g CTAB/L, 1.4 M NaCl, 0.1 M Tris–HCl, 20 mM EDTA) and 1.5uL of MS2 stock were added to a 2mL Precellys SK38 bead beating tube containing a glass/ceramic bead mix, and bead-beat for 2 x 30 seconds at 6.5 m/s using an MP Bio FastPrep Homogeniser. Tubes were centrifuged and the entire sample was transferred to a pre-prepared 1.5mL tube containing 20µL of Proteinase K. Samples were incubated at 65°C for 1 hour. Following centrifugation at 10,000 x g for 15 minutes, 500µL of the supernatant was transferred to a fresh tube and 400µL of chloroform was added, vortexed vigorously, and centrifuged at 10,000xg for 10 minutes. The upper aqueous phase was transferred to a pre-prepared 2mL tube containing a double volume of CTAB precipitation solution (5 g/L, 0.04 M NaCl) and incubated for 1 hour at room temperature. After centrifugation at 16,000 x g for 10 minutes, the supernatant was discarded, and the precipitate was dissolved in 700µL 1.2M NaCl and extracted with an equal volume of chloroform. The mixture was centrifuged for a further 10 minutes at 16,000 x g and the upper aqueous phase was transferred to a fresh tube containing 0.6 volume parts of isopropanol. After overnight incubation at –20°C, the samples were centrifuged at 16,000 x g for 10 minutes and the supernatant was discarded. The pellet was washed with 1mL of 70% ethanol and centrifuged at 16,000 x g for 10 minutes. The supernatant was discarded, and the pellet was dried at 37°C for 1 hour. The dry pellet was dissolved in 100µL molecular-grade water and the extracted nucleic acid was stored at –20°C until use.

Extracts were subsequently analysed by custom TaqMan Array card targeting 29 enteric pathogens: adenovirus 40/41, astrovirus, norovirus GI and GII, rotavirus, sapovirus, *Aeromonas*, *Helicobacter pylori*, *Salmonella enterica (including serovar Typhi), Campylobacter jejuni/coli*, *Shigella* /enteroinvasive *E. coli* (EIEC) and *S. flexneri* and *S. sonnei*, *Clostridium difficile* toxin B, enteroaggregative *E. coli* (EAEC), enteropathogenic *E. coli* (EPEC), enterotoxigenic *E.coli* (ETEC) LT/ST, *E. coli* O157, Shiga-like toxin-producing *E. coli* (STEC) stx1/stx2, *Vibrio cholerae* including toxigenic strains, *Yersinia enterocolitica*, *Plesiomonas shigelloides*, *Entamoeba histolytica*, *Cryptosporidium* spp., *Cyclospora cayetanensis, Ascaris lumbricoides, Trichuris trichiura, Ancylostoma duodenale, Necator americanus, Strongyloides steracoralis*, and *Schistosoma* spp., as well three controls (MS2, bacterial 16S, and a manufacturing control). See Supplemental Table 1 for a detailed list of gene targets and primer and probe sequences. The detection of *Giardia lamblia* was performed using single-tube quantitative PCR with primers and probes, as outlined in Supplemental Table 1.

Sample extracts (40 µL) were combined with 25 µL of TaqPath^TM^ 1-Step RT-qPCR Master Mix (Thermofisher, catalogue # A15300) and 35 µL of molecular grade water before being added to the TAC, centrifuged, sealed, and cycled on QuantStudio 7 Pro as previously described (27). Samples were considered positive for a pathogen if the corresponding gene target(s) had a Cq <35 and it was not detected in the negative extraction control, process control, or no-template control (negative PCR control, run once every 10 cards) (28, 29). Samples were considered negative for a pathogen if the gene target(s) were not detected or had a Cq ≥35, and the extrinsic control (MS2) was detected.

### Data analysis

Descriptive statistics were used to report the characteristics of the study population, prevalence of risk factors, and prevalence of enteric pathogen contamination within food samples. Risk factors explored included sociodemographic characteristics, characteristics related to water, sanitation, and hygiene (WASH), other environmental factors, and types of food consumed. For analysis, child food types were grouped into 4 categories: cereal, mixtures of cereal, meat, fish, and vegetables/legumes, dairy, and other. This grouping was necessary due to the large number of distinct food descriptions with only small counts in each.

To estimate relative wealth, sociodemographic, asset, and wealth variables, commonly used in the Demographic Health Survey (DHS), were used to create a wealth index based on principal component analysis (FactoMineR package: ‘PCA’ function) (30). Five variables were used to construct the index: asset ownership (4 binary variables) and housing characteristics (one binary variable) (Appendix: Table 2).

**Table 1.** Socio-demographic characteristics among caregivers and children in Kenya.

| Variable | Value | N = 569 | (%) or mean (SD) |
| --- | --- | --- | --- |
| **Child level variables** |  |  |  |
| Caregiver gender | Female | 551 | 96.8% |
| Caregiver age | 16-30 | 329 | 57.8% |
|  | 31-45 | 216 | 38.0% |
|  | 46-60 | 21 | 3.7% |
|  | >60 | 3 | 0.5% |
| Caregiver's relationship to infant | Mother | 505 | 88.8% |
|  | Father | 13 | 2.3% |
|  | Grandmother | 27 | 4.7% |
|  | Other | 24 | 4.2% |
| Level of education completed by caregiver | None | 4 | 0.7% |
|  | Primary | 152 | 26.7% |
|  | Secondary | 281 | 49.4% |
|  | Higher | 132 | 23.2% |
| Infant Sex | Female | 285 | 50.1% |
| Infant Age | 6 – 12 months | 179 | 31.5% |
|  | 12 – 18 months | 194 | 34.1% |
|  | 18 – 24 months | 196 | 34.4% |
| Infant ever breastfed | Yes | 562 | 98.8% |
| Infant currently breastfeeds<sup>1</sup> | Yes | 454 | 80.8% |
| **Household level variables** |  |  |  |
| House owned | Yes | 120 | 21.1% |
| Number of household members | 4.4 (1.4) |  |  |
| Number of household members <5 years of age | 1.3 (0.6) |  |  |
| Electricity in household | Yes | 564 | 99.1% |
| Improved material of floor in the house (Cement, mosaic or tile) | Yes | 566 | 99.5% |
| Improved material of walls in the house (Adobe bricks or cement) | Yes | 204 | 35.9% |
| Ownership of livestock (chicken, cow, goat, sheep) | Yes | 106 | 18.6% |
| Human faeces visible on the premises | Yes | 14 | 2.5% |
| Garbage visible on the premises | Yes | 191 | 33.6% |
| Sanitation level (JMP) - At least basic | Yes | 115 | 20.2% |
| Drinking water source (JMP) - At least basic | Yes | 562 | 98.8% |
| Handwashing facility present in household | Yes | 255 | 44.8% |
| Handwashing facility with soap available | Yes | 134 | 23.6% |
| Caregiver reports handwashing in past 24 hours | Yes | 562 | 98.8% |

**Table 2.**
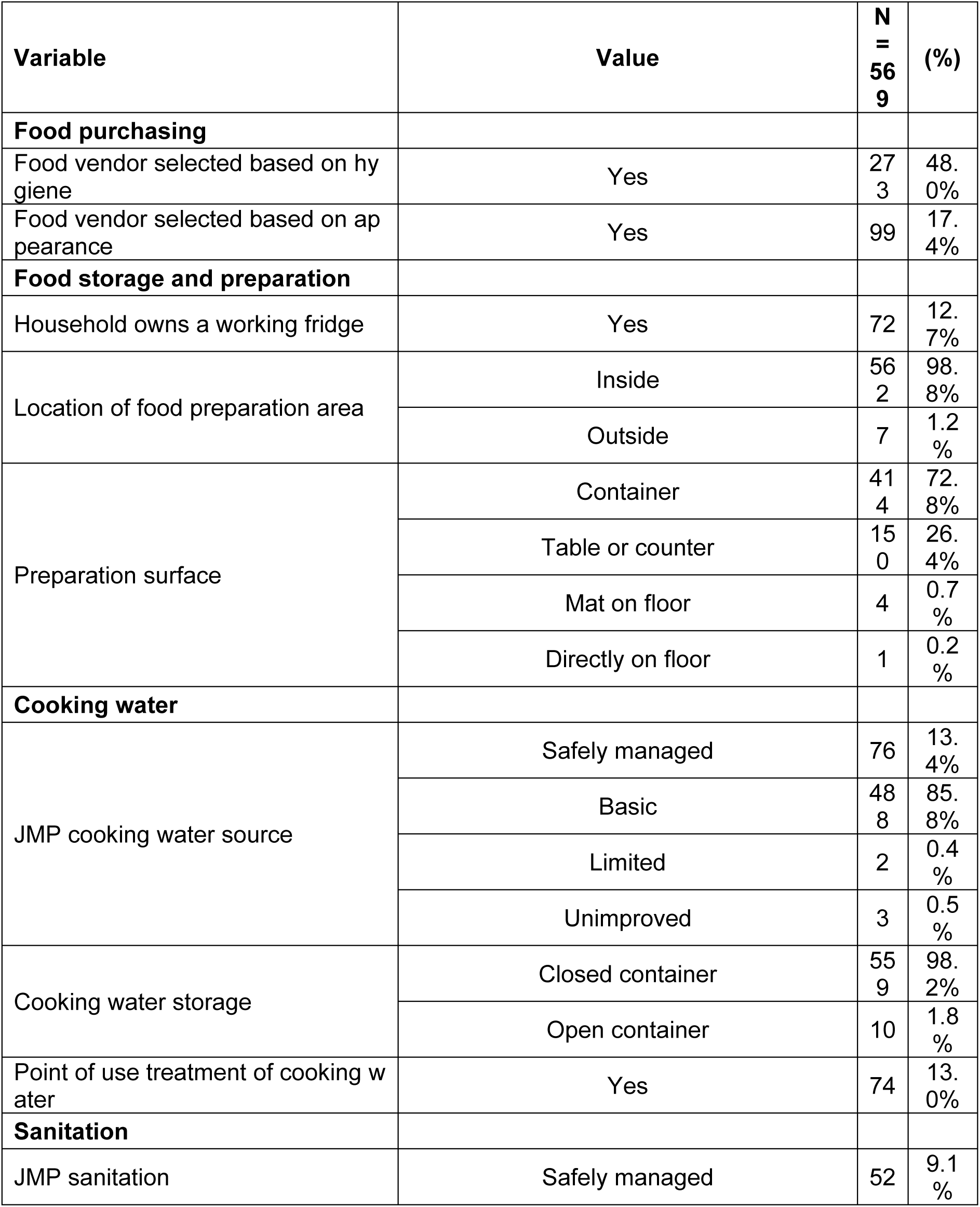
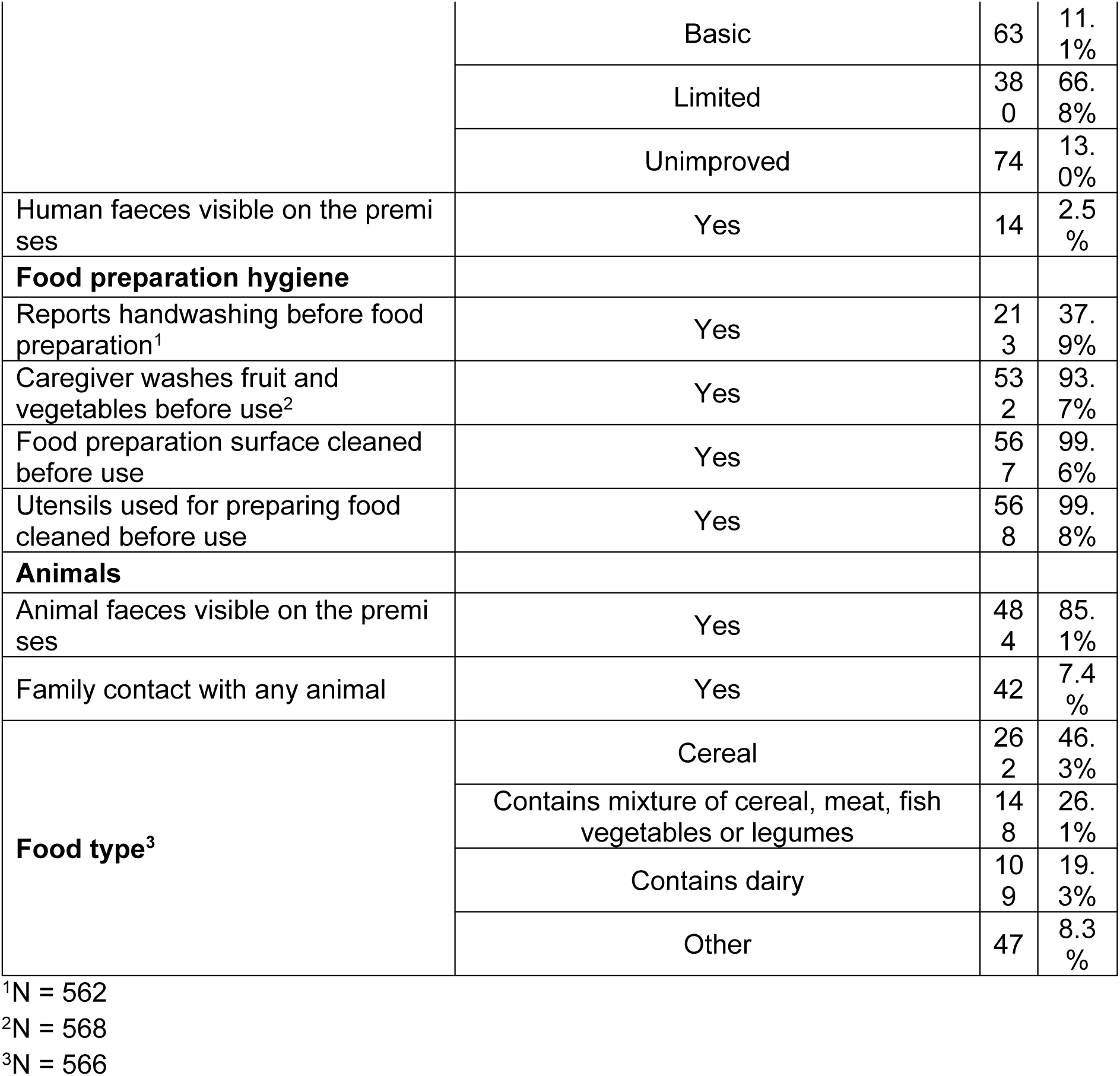
Food-related risk factors among study children in Kenya.

We calculated unadjusted and adjusted odds ratios and 95% confidence intervals to assess each risk factor-outcome relationship and considered the predictor of interest to be significantly associated with the outcome if the p-value was <0.05. Individual risk factors were tested for their odds of association with various outcomes using binomial logistic regression (stats package: ‘glm’ function (31)). Household factors including wealth indices and caregiver education, were selected *a priori* for inclusion in the adjusted multivariable models as potential covariates and confounders. Additional covariates were considered for inclusion in multivariable models if they resulted in a ±10% change in the OR. The final multivariable model was checked for multi-collinearity, with variables exceeding a variance inflation factor of 5 removed (car package: ‘vif’ function (32)). All statistical analyses were conducted in RStudio (33). Given the exploratory nature of these analyses, we did not adjust for multiple comparisons.

### Ethical approval

Ethical approval was obtained from the Research Ethics Committee of the LSHTM (Ref: 17188) and the Institutional Research Ethics Committee at the ILRI (Ref: ILRI-IREC2019-26). The ILRI-IREC is accredited by the National Commission for Science, Technology, and Innovation in Kenya (NACOSTI). In addition, project and individual student approvals were obtained from NACOSTI (License No: NACOSTI/P/21/10409). All study participants (i.e., adult caregivers) provided written informed consent.

## Results

We collected food samples from 569 households in the Dagoretti South Sub-County, Nairobi, Kenya. Cereal was the predominant food type collected (46.3%, n= 262/569) and the remaining samples were grouped into three separate food categories for analysis; a mixture of cereal, meat, fish, vegetables, and legumes (29.3%, n=166/569), dairy (16.1%, n=91/569), and others (8.3%, n=47/569).

### Description of households and study participants

Field teams visited each participating household and collected data on the characteristics of households (Table 1) and food-related risk factors (Table 2). Mothers were the primary caregivers in the households (88.80%, n=505/569) and had completed at least primary education (72.6%, n=413/569). Children were distributed equally between three age groups (<12 months, 12-18 months, and >18 months), and 50.10% (n=285/569) were female. The majority of children were still breastfed (80.80%, n=454/562). Most houses had electricity (99.10%, n=564/569) and the houses were mostly adobe brick built (97.70%, n=556/569) with a basic drinking water source (86.80%, n=494/569). Water was mostly stored in closed containers (98.90%, n=563/569), and most households (66.8%, n=380/569) used limited sanitation facilities (improved facilities shared between two or more households). More than half of all households lacked a handwashing facility (52.5%, n=314/569), and the remaining households used a mobile object for handwashing (30.6%, n=174/569) or a fixed handwashing facility either in the household or inside the compound or yard (15.2%, n=81/569). Only half of those responding reported having soap available (52.50%, n=134/255). Ownership of livestock or companion animals was relatively low (18.60%, n=106/569 and 20.60%, n=117/569, respectively), but 85.10% (n=484/569) of participants reported having contact with animals.

### Food preparation and hygiene

Food hygiene variables relating to infrastructure included the presence of a working fridge (12.70%, n=72/569) to ensure cold storage of food, the presence of a basic Joint Monitoring Programme (JMP) cooking water source (85.80%, n=488/569), (table 2), closed containers for storing cooking water (98.20%, n=559/569), and an inside area for food preparation (98.80%, 562/569). Hygiene behaviours related to food included food vendor selection based on hygiene (48%, n=273/569), handwashing before food preparation (37.90%, 213/562), washing fruit and vegetables before consumption (93.70%, 532.568), cleaning utensils and preparation surfaces (99.80%, n=568/569 and 99.60%, 567/569, respectively), and preparing food inside a container (72.80%, n=414/569) or on a table or counter (26.40%, n=150/469).

### Pathogen prevalence in food

We detected one or more enteric pathogens in 21.4% (n=122/569) of food samples and a mean of 0.3 pathogens per food sample (Table 3). Multiple pathogens were co-detected in 26.3% (n=32/122) of positive food samples. We detected a wide range of pathogens, with 16/30 of the individual enteric pathogens detected in one or more samples. Bacteria were the most detected pathogen type (13.70%, n=78/569), followed by protozoa (9.8%, n=56/569). Viruses (0.40%) and helminths (0.50%) were infrequently detected. Aeromonas (9%, n=51/569), Enterotoxigenic *E. coli* (2.60%, n=15/569), and *Plesiomonas shigelloides* (4.40%, n=25/569) were the most prevalent bacterial pathogens detected, and *Cryptosporidium* (5.30%, n=30/569) and *Giardia* (4.2%, 24/569) were most commonly detected among protozoa (Table 3).

**Table 3.** Detection prevalence of individual and grouped pathogens in food samples.

| Enteric infection | Overall |  |
| --- | --- | --- |
|  | N = 569 | % |
| **Any pathogen** | 122 | 21.40% |
| **Co-detection ($\geq 2$)** | 32 | 5.60% |
| **# of pathogens, Mean (SD)** | 0.3 (0.8) | 0.3 (0.8) |
| **Any virus** | 2 | 0.4% |
| Adenovirus | 0 | 0% |
| Astrovirus | 0 | 0% |
| Norovirus GI/GII | 1 | 0.2% |
| Rotavirus | 1 | 0.2% |
| Sapovirus | 0 | 0% |
| **Any bacteria** | 78 | 13.7% |
| Enteroaggregative *E. coli* | 5 | 0.9% |
| Shigatoxin-producing *E. coli* | 4 | 0.7% |
| *E. coli* O157 | 5 | 0.9% |
| Enteropathogenic *E. coli* | 5 | 0.9% |
| Enterotoxigenic *E. coli* | 15 | 2.6% |
| EIEC/*Shigella* | 1 | 0.2% |
| *Shigella sonnei* | 0 | 0% |
| *Shigella flexneri* | 1 | 0.2% |
| *Campylobacter jejuni/coli* | 0 | 0% |
| *Salmonella enterica* | 0 | 0% |
| *Salmonella typhi* | 0 | 0% |
| Toxigenic *V. cholerae* | 0 | 0% |
| *Clostridium difficile* | 0 | 0% |
| *Yersinia enterocolitica* | 3 | 0.5% |
| *Aeromonas* | 51 | 9.0% |
| *Helicobacter pylori* | 0 | 0% |
| *Plesiomonas shigelloides* | 25 | 4.4% |
| **Any protozoa** | 56 | 9.8% |
| *Cryptosporidium* | 30 | 5.3% |
| *Giardia* | 24 | 4.2% |
| *E. histolytica* | 0 | 0% |
| *Cyclospora cayetanensis* | 5 | 0.9% |
| **Any helminth** | 3 | 0.5% |
| *Ascaris lumbricoides* | 0 | 0% |
| *Trichuris* | 0 | 0% |
| Hookworm | 0 | 0% |
| *Strongyloides stercoralis* | 2 | 0.4% |
| *Schistosoma spp.* | 1 | 0.2% |

### Risk factors for pathogen detection in food

Few food-related exposure variables were significantly associated with the odds of enteric pathogen detection in food (Table 4). We considered the following combined outcomes with a prevalence above 5%: any enteric pathogen, any bacterial pathogen, and any protozoal pathogen detected in food. We also explored the relationship between each risk factor and individual pathogens detected in food with a prevalence above 5%, including *Aeromonas*, *Cryptosporidium*, and any *E. coli*.

**Table 4.** Adjusted regression models for food-related risk factors for the outcomes any pathogen, any bacteria, and any protozoa detection in child food in Kenya.

| | $\geq 1$<br>pathogen | $\geq 1$<br>bacteria | $\geq 1$<br>protozoa | *Aeromonas* | *Cryptosporidium* | $\geq 1$ *E. coli* |
| --- | --- | --- | --- | --- | --- | --- |
| Variable | adjusted OR (95% Confidence interval), p-value |  |  |  |  |  |
| **Food purchasing** |  |  |  |  |  |  |
| Food vendor selected based on hygiene | 0.69<br>(0.46, 1.05),<br>$p=0.084$ | 0.70<br>(0.43, 1.15),<br>$p=0.160$ | 0.55<br>(0.30, 0.99),<br>$p=0.049$ | 0.70<br>(0.38, 1.27),<br>$p=0.245$ | 0.56<br>(0.24, 1.21),<br>$p=0.150$ | 1.35<br>(0.64, 2.93),<br>$p=0.435$ |
| Food vendor selected based on appearance | 0.69<br>(0.38, 1.20),<br>$p=0.209$ | 0.74<br>(0.36, 1.41),<br>$p=0.384$ | 0.71<br>(0.30, 1.50),<br>$p=0.402$ | 0.98<br>(0.43, 2.01),<br>$p=0.955$ | 0.69<br>(0.20, 1.83),<br>$p=0.496$ | 0.51<br>(0.12, 1.49),<br>$p=0.276$ |
| **Food storage and preparation** |  |  |  |  |  |  |
| Household owns a working refrigerator | 0.67<br>(0.32, 1.31),<br>$p=0.262$ | 0.95<br>(0.42, 1.98),<br>$p=0.896$ | 0.71<br>(0.23, 1.86),<br>$p=0.523$<sup>4</sup> | 1.34<br>(0.50, 3.19),<br>$p=0.534$ | 1.62<br>(0.43, 4.97),<br>$p=0.425$<sup>6</sup> | 1.04<br>(0.31, 2.98),<br>$p=0.942$<sup>6</sup> |
| Container used as a preparation surface | 0.75<br>(0.48, 1.18),<br>$p=0.205$ | 0.56<br>(0.33, 0.96),<br>$p=0.033$<sup>5</sup> | 0.73<br>(0.38, 1.45),<br>$p=0.357$<sup>7</sup> | 0.37<br>(0.19, 0.71),<br>$p=0.003$<sup>5,7</sup> | 0.57<br>(0.26, 1.29),<br>$p=0.168$<sup>5</sup> | 0.64<br>(0.30, 1.45),<br>$p=0.266$ |
| **Cooking water** |  |  |  |  |  |  |

|  | ≥1<br>pathogen | ≥1<br>bacteria | ≥1<br>protozoa | Aeromonas | Cryptosporidium | ≥1 E. coli |
| --- | --- | --- | --- | --- | --- | --- |
| Variable | adjusted OR (95% Confidence interval), p-value |  |  |  |  |  |
| JMP cooking water source - safely managed (Ref: Basic, limited, unimproved) | 0.99<br>(0.50, 1.89),<br>p=0.987<sup>4</sup> | 0.76<br>(0.32, 1.64),<br>p=0.500<sup>4</sup> | 0.72<br>(0.24, 1.87),<br>p=0.530<sup>4</sup> | 1.02<br>(0.36, 2.55),<br>p=0.976<sup>4</sup> | 0.87<br>(0.18, 3.12),<br>p=0.844<sup>4,5,6</sup> | 0.36<br>(0.07, 1.29),<br>p=0.151<sup>4,6</sup> |
| Point-of-use treatment of cooking water | 1.47<br>(0.82, 2.56),<br>p=0.186 | 1.74<br>(0.89, 3.23),<br>p=0.089 | 1.25<br>(0.52, 2.70),<br>p=0.587 | 1.64<br>(0.71, 3.46),<br>p=0.215 | 0.82<br>(0.19, 2.47),<br>p=0.759 | 2.40<br>(0.95, 5.56),<br>p=0.048 |
| **Sanitation** |  |  |  |  |  |  |
| JMP sanitation - at least basic (Ref: Limited and unimproved) | 0.86<br>(0.47, 1.55),<br>p=0.629<sup>4</sup> | 1.08<br>(0.55, 2.04),<br>p=0.820 | 0.69<br>(0.28, 1.60),<br>p=0.409<sup>4</sup> | 0.96<br>(0.39, 2.19),<br>p=0.923<sup>4</sup> | 0.68<br>(0.18, 2.15),<br>p=0.544<sup>4,6</sup> | 0.63<br>(0.19, 1.88),<br>p=0.427<sup>4,6,8</sup> |
| Human faeces are visible on the premises | 0.95<br>(0.21, 3.16),<br>p=0.939 | 1.07<br>(0.16, 4.06),<br>p=0.935 | 1.33<br>(0.20, 5.24),<br>p=0.721 | 0.75<br>(0.04, 3.98),<br>p=0.789 | 1.39<br>(0.07, 7.82),<br>p=0.757<sup>6</sup> | NA |
| **Food preparation hygiene** |  |  |  |  |  |  |
| Reports handwashing before food preparation | 0.98<br>(0.64, 1.49),<br>p=0.913 | 0.90<br>(0.54, 1.49),<br>p=0.696 | 1.43<br>(0.80, 2.53),<br>p=0.224 | 0.96<br>(0.51, 1.75),<br>p=0.897 | 1.40<br>(0.65, 2.96),<br>p=0.385 | 1.05<br>(0.46, 2.27),<br>p=0.908 |
| Caregiver washes fruit and vegetables before use<sup>2</sup> | 1.90<br>(0.78, 5.71),<br>p=0.199 | 2.97<br>(0.87, 18.61),<br>p=0.142 | 0.76<br>(0.30, 2.33),<br>p=0.589 | 4.10<br>(0.85, 73.99),<br>p=0.170 | 2.37<br>(0.47, 43.08),<br>p=0.407 | 1.77<br>(0.35, 32.35),<br>p=0.583<sup>8</sup> |
| **Animals** |  |  |  |  |  |  |
| Family contact with any animal | 1.11<br>(0.63, 2.03),<br>p=0.732 | 0.81<br>(0.41, 1.69),<br>p=0.551<sup>6</sup> | 1.56<br>(0.69, 4.21),<br>p=0.324 | 1.36<br>(0.60, 3.67),<br>p=0.494 | 7.34<br>(1.51, 132.30),<br>p=0.053<sup>6</sup> | 0.31<br>(0.11, 0.90),<br>p=0.025<sup>6,8</sup> |
| Animal faeces visible on the premises | 1.60<br>(0.76, 3.20),<br>p=0.197 | 1.90<br>(0.82, 4.05),<br>p=0.111 | 0.99<br>(0.29, 2.66),<br>p=0.992 | 1.44<br>(0.47, 3.60),<br>p=0.474 | 1.16<br>(0.18, 4.36),<br>p=0.846<sup>6</sup> | 1.33<br>(0.30, 4.18),<br>p=0.657<sup>6</sup> |
| Rodents sighted in the household in the past 7 days | 0.91<br>(0.59, 1.40),<br>p=0.660 | 0.60<br>(0.36, 0.99),<br>p=0.044 | 1.55<br>(0.84, 2.98),<br>p=0.174 | 0.58<br>(0.32, 1.07),<br>p=0.077 | 2.64<br>(1.10, 7.37),<br>p=0.042 | 0.41<br>(0.18, 0.88),<br>p=0.023 |
| **Food type<sup>3</sup>** |  |  |  |  |  |  |
| Cereal |  |  |  |  |  |  |
| Contains mixture of cereal, meat, fish, vegetables or legumes | 1.17<br>(0.73, 1.86),<br>p=0.498 | 1.69<br>(0.97, 2.94),<br>p=0.063 | 0.91<br>(0.48, 1.67),<br>p=0.766 | 2.25<br>(1.18, 4.35),<br>p=0.015 | 1.03<br>(0.45, 2.24),<br>p=0.939 | 1.96<br>(0.80, 4.86),<br>p=0.136 |
| Variable | adjusted OR (95% Confidence interval), p-value |  |  |  |  |  |
| Contains dairy | 0.42<br>(0.21,<br>0.78),<br>p=0.009 | 0.94<br>(0.45,<br>1.83),<br>p=0.852 | 0.06<br>(0.00,<br>0.29),<br>p=0.006 | 0.61<br>(0.20,<br>1.56),<br>p=0.335 | NA | 2.16<br>(0.83,<br>5.56),<br>p=0.108 |
| Other | 0.37<br>(0.12,<br>0.90),<br>p=0.046 | 0.47<br>(0.11,<br>1.40),<br>p=0.232 | 0.14<br>(0.01,<br>0.67),<br>p=0.054 | 0.79<br>(0.18,<br>2.48),<br>p=0.720 | 0.28<br>(0.02,<br>1.46),<br>p=0.230 | NA |

<sup>1</sup> N = 562
<sup>2</sup> N = 568
<sup>3</sup> N = 566
<sup>4</sup> Adjusted for WHO water per person
<sup>5</sup> Presence of soap
<sup>6</sup> Ownership of any animal
<sup>7</sup> Handwashing facility present
<sup>8</sup> Over 4 household members
Note: All multivariate models were adjusted for the *a priori* confounders/covariates wealth index and education (higher than primary education)

Caregivers selecting food vendors based on hygiene was found to be associated with decreased odds of detection of ≥1 protozoan pathogen in food (adjusted odds ratio [OR] 0.55, 95% confidence interval [CI] 0.30, 0.99, p= 0.049) (Table 4). Although not significant, this was also associated with decreased odds of ≥1 enteric pathogen detection (adjusted odds ratio [OR] 0.69, 95% confidence interval [CI] 0.46, 1.05, p= 0.084). Preparing food in a container (versus directly on the table/counter) was consistently associated with decreased pathogen detection, although only significant for bacteria (adjusted odds ratio [OR] 0.56, 95% confidence interval [CI] 0.33, 0.96, p= 0.033). For bacteria, the association was most likely driven by *Aeromonas* (adjusted odds ratio [OR] 0.37, 95% confidence interval [CI] 0.19, 0.71, p= 0.003) (Table 4).

Contact with animals was weakly associated with increased odds of detecting C*ryptosporidium* (adjusted odds ratio [OR] 7.34, 95% confidence interval [CI] 1.51-132.30, p= 0.053) (Table 4). Rodent sightings in the household within the last week were associated with increased odds of detecting *Cryptosporidium* (adjusted odds ratio [OR] 2.64, 95% confidence interval [CI] 1.10, 7.37, p=0.042). Conversely, rodent sightings were associated with a reduced risk of detecting bacteria in food (adjusted odds ratio [OR] 0.60, 95% confidence interval [CI] 0.36, 0.99, p=0.044. We created a composite variable of all *E. coli-*positive food samples. Point-of-use treatment of cooking water was associated with increased odds of *E. coli* detection (adjusted odds ratio [OR]2.4, 95% confidence interval [CI] 0.95, 5.56, p= 0.048) and contact with animals was associated with decreased odds of *E. coli* detection once we adjusted for number of household members and animal ownership (unadjusted odds ratio [uOR] 0.56, 95% confidence interval [CI] 0.24, 1.44, p= 0.191, and adjusted odds ratio [OR] 0.31, 95% confidence interval [CI] 0.11, 0.9, p=0.025.

Mixtures of cereal, meat, fish, vegetables, and legumes were weakly associated with increased odds of detecting any bacteria (adjusted odds ratio [OR] 1.69, 95% confidence interval [CI] 0.97, 2.94, p=0.063) in the sample. This relationship was pronounced in the detection of *Aeromonas* (adjusted odds ratio [OR] 2.25, 95% confidence interval [CI] 1.18, 4.35, p=0.015). Foods containing dairy were associated with decreased odds of detecting ≥1 enteric pathogen in the sample (adjusted odds ratio [OR] 0.42, 95% confidence interval [CI] 0.21, 0.78, p=0.009) and ≥1 protozoan parasite (adjusted odds ratio [OR] 0.06, 95% confidence interval [CI] 0, 0.29, p=0.006) even though the number of pathogen-positive dairy samples was low (11.9%, n=13/109) (Supplemental table 2. Results for unadjusted univariable models were similar for all analyses (Supplementary Tables 2 – 7).

## Discussion

We assessed child food contamination in 569 low-income households in the peri-urban Dagoretti South Sub-County, Nairobi, Kenya, and found enteric pathogen contamination in one-fifth of collected food samples. Bacteria were the predominant pathogen type, followed by protozoa. Viruses and helminths were rarely detected. We found that food vendor selection based on hygiene was protective for enteric pathogen detection and more specifically for protozoa detection in food. Preparing food in a container was protective for the detection of bacteria and, more specifically, for *Aeromonas spp*. in food. Contact with animals and rodent sightings in households were associated with increased odds of detecting *Cryptosporidium* in food. In the food type analysis, we found that foods containing dairy were associated with decreased odds of pathogen detection, and mixtures of food containing cereal, meat, fish, vegetables, or legumes were consistently associated with increased odds of detecting bacteria, particularly *Aeromonas*.

In our study, where households selected food vendors based on hygiene and prepared food in a container, we consistently detected an association with decreased odds of enteric pathogen detection in food. In settings where limited resources often restrict the implementation of comprehensive food safety measures, targeted interventions that address food preparation practices and vendor selection present a feasible and effective approach to reducing contamination in food. Previous studies (18, 34) have demonstrated that food safety outcomes improve when vendors are educated in basic hygiene practices, including handwashing, proper food storage, and avoidance of cross-contamination (35). Hygiene certification programs and food hygiene training can be included in public health initiatives to promote better food-handling practices across communities (13). Preparing weaning food in containers helps to prevent exposure of food to environmental contaminants, such as dust, insects, and microorganisms (36). Sealing and cleaning containers before use adds further protection and decreases the risk of pathogen transmission to infants (37–39). In our study site, participants were handed out plastic containers for food storage as part of a different pilot project. We observed that participants decided to repurpose the containers for food preparation without the input from the study team. A recent systematic review suggests that food hygiene interventions focusing on a single intervention can effectively improve the microbial quality of food (15) despite the agreement that food safety needs to be managed from farm to fork. Single interventions, when implemented in low-resource settings, are more likely to promote long-term behavioural change due to their simplicity (40). Single, straightforward interventions are feasible and cost-effective as they require little infrastructure and limited financial investment, and they may also achieve more sustained changes in food hygiene behaviours as they are less challenging to implement (41).

In this study, we found infant food containing a mixture of cereal, meat, fish, vegetables, or legumes was associated with *Aeromonas* detection. The occurrence of *Aeromonas* in these food types is well-documented (42) and further underlines the need for studies on this emerging foodborne pathogen. *Aeromonas spp*. was the most detected pathogen in food in our analysis. The genus consists of facultative Gram-negative bacteria ubiquitously found in aquatic environments (43). In the only other study using TAC to analyse food samples, also set in Kenya, *Aeromonas* was the most prevalent organism (2). *Aeromonas* is an emerging foodborne pathogen, and its role as a true enteric pathogen is still controversial, even though recent studies indicate that they are highly infectious and that human consumption of contaminated food or water results in a high likelihood of disease (44). Emerging antibiotic resistance in *Aeromonas* further warrants continuous monitoring of AMR caused by *Aeromonas* circulating in the food chain and potentially spreading AMR genes to other bacteria inhabiting the same environment or to humans (45, 46).

Animal ownership is often linked to an increase in a household’s exposure to enteric pathogens, particularly if the animals are kept within the household or compound (47). We found household member contacts with animals were associated with a seven-fold increased odds of *Cryptosporidium* contamination of food, in line with previous studies exploring the risk factors for *Cryptosporidium* infection (48–51). The second most detected pathogen in our analysis was *Cryptosporidium*, consistent with a previous study (2). As cryptosporidiosis is the second-highest cause of moderate-to-severe diarrhoea in under-5-year-olds (9) interrupting the diverse pathways of *Cryptosporidium* spread is important in the prevention of severe foodborne outbreaks (52). A variety of *Cryptosporidium* species and genotypes, encompassing a wide host range, present a significant threat to food safety through contamination of the food and water supply (49, 51). Due to their zoonotic and anthroponotic potential, these protozoan parasites represent a growing burden on both the public health and veterinary sectors (48). The high incidence of foodborne and waterborne outbreaks observed not only in low-and middle-income countries but also in high-income nations underscores the urgent need for effective control measures against *Cryptosporidium*.

Conversely, animal contact was found to be protective of food contamination with *E. coli* in our analysis. In a multi-site study in Mali and South Sudan (53), animal contact was found to be protective for relapse to acute malnutrition, likely due to the confounding effect of animal ownership association with wealth in more rural areas. Contrary to other studies, dairy was protective of pathogen detection in infant food (2, 17). In another study in a similar setting in Kisumu, Kenya, cow’s milk was significantly more likely to be contaminated with enteric pathogens compared to other common infant foods, such as porridge (2). Our analysis was most likely not powered to detect an association between dairy and pathogen detection, considering the very low number of pathogen-positive dairy samples, and the protective effect of dairy may reflect differences in the cooking, handling, or storage of dairy products relative to other foods in our study area.

One of the strengths of our study lies in the direct examination of food samples for pathogens, rather than using faecal indicators as a proxy for assessing the risk of faecal pathogen detection. Real-time quantitative PCR (qPCR) is routinely used in foodborne pathogen outbreaks in HICs, but the use of qPCR in epidemiological studies in LMICs is still limited (54, 55). Further, traditional qPCR tests for 1-6 targets in a single reaction. The microfluidic technology of TAC used here allows for the simultaneous detection of 48 different targets, thereby enabling the analysis of small volumes of precious samples (56). Our study had several limitations; due to the cross-sectional nature of the study, we were unable to establish a causal relationship between the detection of enteric pathogens in food and diarrhoeal disease in children. In another study in the same study area, Knee *et al* confirmed high levels of enteric pathogen exposure among children, exploring the same pathogens in stool (25). Due to the limited variability in responses for certain risk factors and the low prevalence of detection for specific outcomes (e.g., helminths and viruses), it was not possible to assess the relationships between all food risk factors and outcomes. In addition, the number of risk factors and outcomes analysed could also mean that some of the findings were spurious and due to chance.

We showed that complementary foods collected from households in this setting carry a variety of enteric pathogens. Contaminated food is an important pathway of disease transmission in infants. Targeted food hygiene interventions are essential to interrupt disease transmission and to reduce the exposure of children to enteric pathogens, particularly during vulnerable weaning periods. Our analysis identified potential behavioural changes in domestic food practices that may lead to reduced food contamination. Future research should focus on longitudinal studies elucidating the impact of simple control measures, such as minimizing animal contact within households, improving food preparation practices, interventions at the vendor level, and modifying food purchasing behaviours on the foodborne transmission of pathogens in infants.

## Data Availability

All data produced in the present study are available upon reasonable request to the authors

## Acknowledgements

We would like to extend our deep gratitude to the residents and communities of Dagoretti for their participation in this study. We would also like to thank all members of the field research, laboratory, and data management teams for their continued support throughout the project.

