## Supplementary Data for "Risk factors for enteric pathogen detection in food consumed by children aged 6–24 months in informal urban neighbourhoods of Nairobi, Kenya: a cross-sectional study"

**Supplemental Table 1**: TaqMan Array Card molecular gene targets and sequences

| Category | Pathogen | Target | Primers & probes sequences  (labelled with FAM (6-carboxyfluorescein) at 5′ and MGB at 3′) |
| --- | --- | --- | --- |
| Virus | Adenovirus F (40/41) | fiber gene | F: AACTTTCTCTCTTAATAGACGCC; R: AGGGGGCTAGAAAACAAAA  Probe: CTGACACGGGCACTCT |
|  | Astrovirus | Capsid | F: CAGTTGCTTGCTGCGTTCA; R: CTTGCTAGCCATCACACTTCT  Probe: CACAGAAGAGCAACTCCATCGC |
|  | Norovirus GI | ORF1-2 | F: CGYTGGATGCGNTTYCATGA; R: CTTAGACGCCATCATCATTYAC  Probe: TGGACAGGAGATCGC |
|  | Norovirus GII | ORF1-2 | F: CARGARBCNATGTTYAGRTGGATGAG; R: TCGACGCCATCTTCATTCACA  Probe: TGGGAGGGCGATCGCAATCT |
|  | Rotavirus | NSP3 | F: ACCATCTWCACRTRACCCTCTATGAG; R: GGTCACATAACGCCCCTATAGC  Probe: AGTTAAAAGCTAACACTGTCAAA |
|  | Sapovirus | RdRp | Fw1: GAYCASGCTCTCGCYACCTAC; Fw2: TTGGCCCTCGCCACCTAC;  R: CCCTCCATYTCAAACACTA  Probe: CCRCCTATRAACCA |
| Bacteria | Aeromonas | aerolysin | F: TYCGYTACCAGTGGGACAAG; R: CCRGCAAACTGGCTCTCG  Probe: CAGTTCCAGTCCCACCACTT |
|  | Campylobacter jejuni/coli | cadF | F: CTGCTAAACCATAGAAATAAAATTTCTCAC; R: CTTTGAAGGTAATTTAGATATGGATAATCG  Probe: CATTTTGACGATTTTTGGCTTGA |
|  | C. difficile | tcdB | F: GGTATTACCTAATGCTCCAAATAG; R: TTTGTGCCATCATTTTCTAAGC  Probe: CCTGGTGTCCATCCTGTTTC |
|  | EAEC | aaiC | F: ATTGTCCTCAGGCATTTCAC; R: ACGACACCCCTGATAAACAA  Probe: TAGTGCATACTCATCATTTAAG |
|  | EAEC | aatA | F: CTGGCGAAAGACTGTATCAT; R: TTTTGCTTCATAAGCCGATAGA  Probe: TGGTTCTCATCTATTACAGACAGC |
|  | EAEC | aggR | F: GCAATCAGATTAARCAGCGATACA; R: TTCGGACAACTRCAAGCATC  Probe: AAGACGCCTAAAGGATGCCC |
|  | STEC | stx1 | F: ACTTCTCGACTGCAAAGACGTATG; R: ACAAATTATCCCCTGWGCCACTATC  Probe: CTCTGCAATAGGTACTCCA |
|  | STEC | stx2 | F: CCACATCGGTGTCTGTTATTAACC; R: GGTCAAAACGCGCCTGATAG  Probe: TTGCTGTGGATATACGAGG |
|  | EPEC | Eae | F: CATTGATCAGGATTTTTCTGGTGATA; R: CTCATGCGGAAATAGCCGTTA  Probe: ATACTGGCGAGACTATTTCAA |
|  | EPEC | bfpA | F: TGGTGCTTGCGCTTGCT; R: CGTTGCGCTCATTACTTCTG  Probe: CAGTCTGCGTCTGATTCCAA |
|  | ETEC | LT | F: TTCCCACCGGATCACCAA; R: CAACCTTGTGGTGCATGATGA  Probe: CTTGGAGAGAAGAACCCT |
|  | ETEC | STh | F: GCTAAACCAGYAGRGTCTTCAAAA; R: CCCGGTACARGCAGGATTACAACA  Probe: TGGTCCTGAAAGCATGAA |
|  | ETEC | STp | F: TGAATCACTTGACTCTTCAAAA; R: GGCAGGATTACAACAAAGTT  Probe: TGAACAACACATTTTACTGCT |
|  | E. coli O157 | rfbE | F: TTTCACACTTATTGGATGGTCTCAA; R: CGATGAGTTTATCTGCAAGGTGAT  Probe: CTCTCTTTCCTCTGCGGTCCT |
|  | Helicobacter pylori | ureC | F: GACACCAGAAAAAGCGGCTA; R: AGCGCATGTCTTCGGTTAAA  Probe: TCACTAAAGCGTTTTCTACC |
|  | Plesiomonas shigelloides | gyrB | F: CCGCCGTGAAGGCAAAG; R: GCTACCGGCTCACCCAGAT  Probe: CACACCCAAGAATAC |
|  | Salmonella enterica | ttr | F: CTCACCAGGAGATTACAACATGG; R: AGCTCAGACCAAAAGTGACCATC  Probe: CACCGACGGCGAGACCGACTTT |
|  | Salmonella enterica Typhi | STY0201 | F: CGCGAAGTCAGAGTCGACATAG; R: AAGACCTCAACGCCGATCAC  Probe: CAGCCTGCTCCAGAACA |
|  | Shigella/EIEC | ipaH | F: CCTTTTCCGCGTTCCTTGA; R: CGGAATCCGGAGGTATTGC  Probe: CGCCTTTCCGATACCGTCTCTGCA |
|  | Shigella flexneri | Putative periplasmic  Protein* | F: TGGGTGCATCCTGACCTGT; R: GACAAACAATAACGAGCTACCGAT  Probe: ACCACGGAATAATCCCGCAG |
|  | Shigella flexneri | O-antigen** | F: CTCCTATCCGTGATTATAGTGCA; R: GCACACACAACTCACTGTATTT  Probe: TCCTTCTCACGATTAAAATC |
|  | Shigella flexneri | Type 3 restriction  Enzyme** | F: CTTTCAACGCACGAATATCAAC; R: GAACCTGATCCAGACGGAGA  Probe: TTCTTCAGAACCGGGTTTTG |
|  | Shigella sonnei | Putative methylase | F: TGCCGCTAAAATCCTTCTGT; R: GCGTACGACGAAAGGAAAAA  Probe: GAAGTTATTGATTCCGCCC |
|  | Vibrio cholerae | hlyA | F: ATCGTCAGTTTGGAGCCAGT; R: TCGATGCGTTAAACACGAAG  Probe: ACCGATGCGATTGCCCAA |
|  | Vibrio cholerae | ctxA | F: GCATAGAGCTTGGAGGGAAGAG; R: CATCGATGATCTTGGAGCATTC  Probe: CATCATGCACCGCCG |
|  | Yersinia enterocolitica | lytA | F: TGATTCACCAGCAGCAATAC; R: GGCATCATGAAAGGCGG  Probe: TGTCGGTTTCTCCTTCCAGG |
| Protozoa | Cryptosporidium spp. | 18S rRNA | F: GGGTTGTATTTATTAGATAAAGAACCA; R: AGGCCAATACCCTACCGTCT  Probe: TGACATATCATTCAAGTTTCTGAC |
|  | Cyclospora cayetanensis | 18S | F: AAAAGCTCGTAGTTGGATTTCTG; R: AACACCAACGCACGCAGC  P: AAGGCCGGATGACCACGA |
|  | Giardia spp. *** | 18S rRNA | F: ATCCGGTCGATCCTGCCG ; R: GGGGTGCAACCGTTGTCCT  Probe: CGGCGGACGGCTCAGGAC |
|  | E. histolytica | 18S rRNA | F: ATTGTCGTGGCATCCTAACTCA; R: GCGGACGGCTCATTATAACA,  Probe: TCATTGAATGAATTGGCCATTT |
| Helminth | Ascaris lumbricoides | ITS1 | F: GCCACATAGTAAATTGCACACAAAT; R: GCCTTTCTAACAAGCCCAACAT  Probe: TTGGCGGACAATTGCATGCGAT |
|  | Trichuris trichiura | 18S rRNA | F: TTGAAACGACTTGCTCATCAACTT; R: CTGATTCTCCGTTAACCGTTGTC  Probe: CGATGGTACGCTACGTGCTTACCATGG |
|  | Ancylostoma duodenale | ITS2 | F: GAATGACAGCAAACTCGTTGTTG; R: ATACTAGCCACTGCCGAAACGT  Probe: ATCGTTTACCGACTTTAG |
|  | Necator americanus | ITS2 | F: CTGTTTGTCGAACGGTACTTGC; R: ATAACAGCGTGCACATGTTGC  Probe: CTGTACTACGCATTGTATAC |
|  | Strongyloides stercoralis | dispersed repetitive sequence | F: TCCAGAAAAGTCTTCACTCTCCAG; R: TGCGTTAGAATTTAGATATTATTGTTGCT  Probe: TCAGCTCCAGTTGAACAACAGCCTCCAA |
|  | Schistosoma spp. | ITS | F: GGTCTAGATGACTTGATYGAGATGCT; R: TCCCGAGCGYGTATAATGTCATTA  P: TGGGTTGTGCTCGAGTCGTGGC |
| Other/ virus | SARS-CoV-2# | N1 | F: GACCCCAAAATCAGCGAAAT; R: TCTGGTTACTGCCAGTTGAATCTG  Probe: ACCCCGCATTACGTTTGGTGGACC |
| Other/ virus | SARS-CoV-2# | E-Sarbeco | F: ACAGGTACGTTAATAGTTAATAGCGT; R: ATATTGCAGCAGTACGCACACA  Probe: ACACTAGCCATCCTTACTGCGCTTCG |
| Control/ RNA virus | MS2 | MS2g1 | F: TGGCACTACCCCTCTCCGTATTCAC; R: GTACGGGCGACCCCACGATGAC  Probe: CACATCGATAGATCAAGGTGCCTACAAGC |
| Control/ DNA virus | PhHV | gB | F: GGGCGAATCACAGATTGAATC; R: GCGGTTCCAAACGTACCAA  Probe: TATGTGTCCGCCACCATCT |
| Control/ 16S rRNA | 16S | 16S | F: TGCAAGTCGAACGAAGCACTTTA; R: GCAGGTTACCCACGCGTTAC  Probe: CGCCACTCAGTCACAAA |
| Control/ 18S rRNA | 18S† | 18S | Manufacturer's control |

*This assay detects most *S. flexneri* serotypes except for serotype 6.

**The combination of these two assays identifies *S. flexneri* serotype 6 when both are positive (Cq≤35)

****Giardia lamblia* analysed by single tube qPCR. Primer and probe concentration, mastermix, and cycling conditions equivalent to TAC analysis.

\# Included on the TAC but not reported in this manuscript as not a enteric pathogen primarily transmitted via faecal-oral route.

†ThermoFisher manufacturer control

All primer and probe sequences from (27).

|  | | [final] |
| --- | --- | --- |
| TaqPath^TM^ 1-Step RT-qPCR Master Mix | | 1x |
| Primer Fw & Rv (*Giardia*) [each] | | 900 nM |
| *Giardia*_probe (FAM) | | 250 nM |
| ddH2O | | . |
| template | | . |

**Supplementary table 2.** Unadjusted and adjusted regression models for food-related risk factors and their association with any pathogen detection in child food in Kenya.

|  | **Overall, N = 569** | **No enteric detection, N = 447** | **Enteric detection, N = 122** | **Unadjusted** | | | **Adjusted** | | |
| --- | --- | --- | --- | --- | --- | --- | --- | --- | --- |
| **Variable** |  |  |  | **uOR** | **95% CI** | **p-value** | **aOR** | **95% CI** | **p-value** |
| **Food purchasing** |  |  |  |  |  |  |  |  |  |
| Food vendor selected based on hygiene | 273 (48.0%) | 223 (49.9%) | 50 (41.0%) | 0.70 | 0.46, 1.04 | 0.082 | 0.69 | 0.46, 1.05 | 0.084 |
| Food vendor selected based on appearance | 99 (17.4%) | 82 (18.3%) | 17 (13.9%) | 0.72 | 0.40, 1.24 | 0.256 | 0.69 | 0.38, 1.20 | 0.209 |
| **Food storage and preparation** |  |  |  |  |  |  |  |  |  |
| Household owns a working refrigerator | 72 (12.7%) | 60 (13.4%) | 12 (9.8%) | 0.70 | 0.35, 1.31 | 0.293 | 0.67 | 0.32, 1.31 | 0.262 |
| Container used as preparation surface | 414 (72.8%) | 331 (74.0%) | 83 (68.0%) | 0.75 | 0.48, 1.16 | 0.187 | 0.75 | 0.48, 1.18 | 0.205 |
| **Cooking water** |  |  |  |  |  |  |  |  |  |
| JMP cooking water source - safely managed (Ref: Basic, limited, unimproved) | 76 (13.4%) | 58 (13.0%) | 18 (14.8%) | 1.16 | 0.64, 2.02 | 0.609 | 0.99 | 0.50, 1.89 | 0.987 |
| Point of use treatment of cooking water | 74 (13.0%) | 54 (12.1%) | 20 (16.4%) | 1.43 | 0.80, 2.46 | 0.211 | 1.47 | 0.82, 2.56 | 0.186 |
| **Sanitation** |  |  |  |  |  |  |  |  |  |
| JMP sanitation - at least basic (Ref: Limited and unimproved) | 115 (20.2%) | 91 (20.4%) | 24 (19.7%) | 0.96 | 0.57, 1.56 | 0.867 | 0.86 | 0.47, 1.55 | 0.629 |
| Human faeces visible on the premises | 14 (2.5%) | 11 (2.5%) | 3 (2.5%) | 1.00 | 0.22, 3.26 | >0.999 | 0.95 | 0.21, 3.16 | 0.939 |
| **Food preparation hygiene** |  |  |  |  |  |  |  |  |  |
| Reports handwashing before food preparation^1^ | 213 (37.9%) | 169 (38.2%) | 44 (36.7%) | 0.94 | 0.61, 1.42 | 0.753 | 0.98 | 0.64, 1.49 | 0.913 |
| Caregiver washes fruit and vegetables before use^2^ | 532 (93.7%) | 415 (93.0%) | 117 (95.9%) | 1.75 | 0.72, 5.21 | 0.257 | 1.90 | 0.78, 5.71 | 0.199 |
| **Animals** |  |  |  |  |  |  |  |  |  |
| Family contact with any animal | 484 (85.1%) | 379 (84.8%) | 105 (86.1%) | 1.11 | 0.64, 2.02 | 0.726 | 1.11 | 0.63, 2.03 | 0.732 |
| Animal faeces visible on the premises | 42 (7.4%) | 30 (6.7%) | 12 (9.8%) | 1.52 | 0.73, 2.99 | 0.245 | 1.60 | 0.76, 3.20 | 0.197 |
| Rodents sighted in household in past 7 days | 359 (63.1%) | 285 (63.8%) | 74 (60.7%) | 0.88 | 0.58, 1.33 | 0.529 | 0.91 | 0.59, 1.40 | 0.660 |
| **Food type^3^** |  |  |  |  |  |  |  |  |  |
| Cereal | 262 (46.3%) | 200 (44.9%) | 62 (51.2%) | — |  |  |  |  |  |
| Contains mixture of cereal, meat, fish vegetables or legumes | 148 (26.1%) | 110 (23.9%) | 38 (35.8%) | 1.46 | 0.90, 2.37 | 0.120 | 1.40 | 0.86, 2.27 | 0.175 |
| Dairy | 109 (19.3%) | 96 (20.9%) | 13 (12.3%) | 0.57 | 0.29, 1.08 | 0.097 | 0.55 | 0.28, 1.05 | 0.080 |
| Other | 47 (8.3%) | 42 (9.1%) | 5 (4.7%) | 0.50 | 0.17, 1.23 | 0.170 | 0.48 | 0.16, 1.18 | 0.144 |

^1^ N = 562

^2^ N = 568

^3^ N = 566

**Supplementary table 3.** Unadjusted and adjusted regression models for food-related risk factors and their association with any bacteria detection in child food in Kenya.

| **Variable** | **Overall, N = 569** | **No bacteria detection, N = 491** | **Bacteria detection, N = 78** | **Unadjusted** | | | **Adjusted** | | |
| --- | --- | --- | --- | --- | --- | --- | --- | --- | --- |
|  |  |  |  | **uOR** | **95% CI** | **p-value** | **aOR** | **95% CI** | **p-value** |
| **Food purchasing** |  |  |  |  |  |  |  |  |  |
| Food vendor selected based on hygiene | 273 (48.0%) | 241 (49.1%) | 32 (41.0%) | 0.72 | 0.44, 1.17 | 0.187 | 0.70 | 0.43, 1.15 | 0.160 |
| Food vendor selected based on appearance | 99 (17.4%) | 88 (17.9%) | 11 (14.1%) | 0.75 | 0.36, 1.43 | 0.410 | 0.74 | 0.36, 1.41 | 0.384 |
| **Food storage and preparation** |  |  |  |  |  |  |  |  |  |
| Household owns a working refrigerator | 72 (12.7%) | 62 (12.6%) | 10 (12.8%) | 1.02 | 0.47, 2.00 | 0.962 | 0.95 | 0.42, 1.98 | 0.896 |
| Container used as preparation surface | 414 (72.8%) | 364 (74.1%) | 50 (64.1%) | 0.62 | 0.38, 1.04 | 0.066 | 0.56 | 0.33, 0.96 | 0.033 |
| **Cooking water** |  |  |  |  |  |  |  |  |  |
| JMP cooking water source - safely managed (Ref: Basic, limited, unimproved) | 76 (13.4%) | 66 (13.4%) | 10 (12.8%) | 0.95 | 0.44, 1.86 | 0.881 | 0.76 | 0.32, 1.64 | 0.500 |
| Point of use treatment of cooking water | 74 (13.0%) | 59 (12.0%) | 15 (19.2%) | 1.74 | 0.91, 3.19 | 0.082 | 1.74 | 0.89, 3.23 | 0.089 |
| **Sanitation** |  |  |  |  |  |  |  |  |  |
| JMP sanitation - at least basic (Ref: Limited and unimproved) | 115 (20.2%) | 98 (20.0%) | 17 (21.8%) | 1.12 | 0.61, 1.96 | 0.708 | 1.08 | 0.55, 2.04 | 0.820 |
| Human faeces visible on the premises | 14 (2.5%) | 12 (2.4%) | 2 (2.6%) | 1.05 | 0.16, 3.95 | 0.949 | 1.07 | 0.16, 4.06 | 0.935 |
| **Food preparation hygiene** |  |  |  |  |  |  |  |  |  |
| Reports handwashing before food preparation^1^ | 213 (37.9%) | 186 (38.3%) | 27 (35.5%) | 0.89 | 0.53, 1.46 | 0.647 | 0.90 | 0.54, 1.49 | 0.696 |
| Caregiver washes fruit and vegetables before use^2^ | 532 (93.7%) | 456 (93.1%) | 76 (97.4%) | 2.83 | 0.84, 17.7 | 0.158 | 2.97 | 0.87, 18.61 | 0.142 |
| **Animals** |  |  |  |  |  |  |  |  |  |
| Family contact with any animal | 484 (85.1%) | 418 (85.1%) | 66 (84.6%) | 0.96 | 0.51, 1.95 | 0.905 | 0.81 | 0.41, 1.69 | 0.551 |
| Animal faeces visible on the premises | 42 (7.4%) | 33 (6.7%) | 9 (11.5%) | 1.81 | 0.79, 3.80 | 0.136 | 1.90 | 0.82, 4.05 | 0.111 |
| Rodents sighted in household in past 7 days | 359 (63.1%) | 318 (64.8%) | 41 (52.6%) | 0.60 | 0.37, 0.98 | 0.039 | 0.60 | 0.36, 0.99 | 0.044 |
| **Food type^3^** |  |  |  |  |  |  |  |  |  |
| Cereal | 262 (46.3%) | 230 (47.0%) | 32 (41.6%) | — |  |  |  |  |  |
| Contains mixture of cereal, meat, fish vegetables or legumes | 148 (26.1%) | 119 (24.3%) | 29 (37.7%) | 1.75 | 1.01, 3.04 | 0.045 | 1.69 | 0.97, 2.94 | 0.063 |
| Contains dairy | 109 (19.3%) | 96 (19.6%) | 13 (16.9%) | 0.97 | 0.47, 1.90 | 0.938 | 0.94 | 0.45, 1.83 | 0.852 |
| Other | 47 (8.3%) | 44 (9.0%) | 3 (3.9%) | 0.49 | 0.11, 1.45 | 0.254 | 0.47 | 0.11, 1.40 | 0.232 |

^1^ N = 562

^2^ N = 568

^3^ N = 566

**Supplementary table 4.** Unadjusted and adjusted regression models for food-related risk factors and their association with any protozoa detection in child food in Kenya.

| **Variable** | **Overall, N = 569** | **No protozoa detection,**  **N = 513** | **Protozoa detection,**  **N = 56** | **Unadjusted** | | | **Adjusted** | | |
| --- | --- | --- | --- | --- | --- | --- | --- | --- | --- |
|  |  |  |  | **uOR** | **95% CI** | **p-value** | **aOR** | **95% CI** | **p-value** |
| **Food purchasing** |  |  |  |  |  |  |  |  |  |
| Food vendor selected based on hygiene | 273 (48.0%) | 254 (49.5%) | 19 (33.9%) | 0.52 | 0.29, 0.92 | 0.029 | 0.55 | 0.30, 0.99 | 0.049 |
| Food vendor selected based on appearance | 99 (17.4%) | 91 (17.7%) | 8 (14.3%) | 0.77 | 0.33, 1.60 | 0.518 | 0.71 | 0.30, 1.50 | 0.402 |
| **Food storage and preparation** |  |  |  |  |  |  |  |  |  |
| Household owns a working refrigerator | 72 (12.7%) | 67 (13.1%) | 5 (8.9%) | 0.65 | 0.22, 1.55 | 0.380 | 0.71 | 0.23, 1.86 | 0.523 |
| Container used as preparation surface | 414 (72.8%) | 375 (73.1%) | 39 (69.6%) | 0.84 | 0.47, 1.58 | 0.582 | 0.73 | 0.38, 1.45 | 0.357 |
| **Cooking water** |  |  |  |  |  |  |  |  |  |
| JMP cooking water source - safely managed (Ref: Basic, limited, unimproved) | 76 (13.4%) | 70 (13.6%) | 6 (10.7%) | 0.76 | 0.28, 1.71 | 0.542 | 0.72 | 0.24, 1.87 | 0.530 |
| Point of use treatment of cooking water | 74 (13.0%) | 66 (12.9%) | 8 (14.3%) | 1.13 | 0.48, 2.37 | 0.764 | 1.25 | 0.52, 2.70 | 0.587 |
| **Sanitation** |  |  |  |  |  |  |  |  |  |
| JMP sanitation - at least basic (Ref: Limited and unimproved) | 115 (20.2%) | 106 (20.7%) | 9 (16.1%) | 0.74 | 0.33, 1.48 | 0.418 | 0.69 | 0.28, 1.60 | 0.409 |
| Human faeces visible on the premises | 14 (2.5%) | 12 (2.3%) | 2 (3.6%) | 1.55 | 0.24, 5.87 | 0.575 | 1.33 | 0.20, 5.24 | 0.721 |
| **Food preparation hygiene** |  |  |  |  |  |  |  |  |  |
| Reports handwashing before food preparation^1^ | 213 (37.9%) | 189 (37.3%) | 24 (43.6%) | 1.30 | 0.74, 2.28 | 0.357 | 1.43 | 0.80, 2.53 | 0.224 |
| Caregiver washes fruit and vegetables before use^2^ | 532 (93.7%) | 481 (93.9%) | 51 (91.1%) | 0.66 | 0.27, 1.99 | 0.405 | 0.76 | 0.30, 2.33 | 0.589 |
| **Animals** |  |  |  |  |  |  |  |  |  |
| Family contact with any animal | 484 (85.1%) | 434 (84.6%) | 50 (89.3%) | 1.52 | 0.68, 4.06 | 0.353 | 1.56 | 0.69, 4.21 | 0.324 |
| Animal faeces visible on the premises | 42 (7.4%) | 38 (7.4%) | 4 (7.1%) | 0.96 | 0.28, 2.51 | 0.943 | 0.99 | 0.29, 2.66 | 0.992 |
| Rodents sighted in household in past 7 days | 359 (63.1%) | 319 (62.2%) | 40 (71.4%) | 1.52 | 0.84, 2.86 | 0.176 | 1.55 | 0.84, 2.98 | 0.174 |
| **Food type^3^** |  |  |  |  |  |  |  |  |  |
| Cereal | 262 (46.3%) | 228 (44.6%) | 34 (61.8%) | — |  |  |  |  |  |
| Contains mixture of cereal, meat, fish vegetables or legumes | 148 (26.1%) | 129 (25.2%) | 19 (34.5%) | 0.99 | 0.53, 1.79 | 0.968 | 0.91 | 0.48, 1.67 | 0.766 |
| Contains dairy | 109 (19.3%) | 108 (21.1%) | 1 (1.8%) | 0.06 | 0.00, 0.29 | 0.007 | 0.06 | 0.00, 0.29 | 0.006 |
| Other | 47 (8.3%) | 46 (9.0%) | 1 (1.8%) | 0.15 | 0.01, 0.70 | 0.061 | 0.14 | 0.01, 0.67 | 0.054 |

^1^ N = 562

^2^ N = 568

^3^ N = 566

**Supplementary table 5.** Unadjusted and adjusted regression models for food-related risk factors and their association with Aeromonas detection in child food in Kenya.

| **Variable** | **Overall,**  **N = 569** | **No Aeromonas detection,**  **N = 518** | **Aeromonas detection,**  **N = 51** | **Unadjusted** | | | **Adjusted** | | |
| --- | --- | --- | --- | --- | --- | --- | --- | --- | --- |
|  |  |  |  | **uOR** | **95% CI** | **p-value** | **aOR** | **95% CI** | **p-value** |
| **Food purchasing** |  |  |  |  |  |  |  |  |  |
| Food vendor selected based on hygiene | 273 (48.0%) | 253 (48.8%) | 20 (39.2%) | 0.68 | 0.37, 1.21 | 0.191 | 0.70 | 0.38, 1.27 | 0.245 |
| Food vendor selected based on appearance | 99 (17.4%) | 90 (17.4%) | 9 (17.6%) | 1.02 | 0.45, 2.08 | 0.961 | 0.98 | 0.43, 2.01 | 0.955 |
| **Food storage and preparation** |  |  |  |  |  |  |  |  |  |
| Household owns a working refrigerator | 72 (12.7%) | 65 (12.5%) | 7 (13.7%) | 1.11 | 0.44, 2.42 | 0.809 | 1.34 | 0.50, 3.19 | 0.534 |
| Container used as preparation surface | 414 (72.8%) | 385 (74.3%) | 29 (56.9%) | 0.46 | 0.25, 0.83 | 0.009 | 0.37 | 0.19, 0.71 | 0.003 |
| **Cooking water** |  |  |  |  |  |  |  |  |  |
| JMP cooking water source - safely managed (Ref: Basic, limited, unimproved) | 76 (13.4%) | 69 (13.3%) | 7 (13.7%) | 1.04 | 0.41, 2.26 | 0.935 | 1.02 | 0.36, 2.55 | 0.976 |
| Point of use treatment of cooking water | 74 (13.0%) | 65 (12.5%) | 9 (17.6%) | 1.49 | 0.66, 3.08 | 0.304 | 1.64 | 0.71, 3.46 | 0.215 |
| **Sanitation** |  |  |  |  |  |  |  |  |  |
| JMP sanitation - at least basic (Ref: Limited and unimproved) | 115 (20.2%) | 105 (20.3%) | 10 (19.6%) | 0.96 | 0.44, 1.91 | 0.911 | 0.96 | 0.39, 2.19 | 0.923 |
| Human faeces visible on the premises | 14 (2.5%) | 13 (2.5%) | 1 (2.0%) | 0.78 | 0.04, 4.03 | 0.810 | 0.75 | 0.04, 3.98 | 0.789 |
| **Food preparation hygiene** |  |  |  |  |  |  |  |  |  |
| Reports handwashing before food preparation^1^ | 213 (37.9%) | 195 (38.1%) | 18 (36.0%) | 0.91 | 0.49, 1.66 | 0.772 | 0.96 | 0.51, 1.75 | 0.897 |
| Caregiver washes fruit and vegetables before use^2^ | 532 (93.7%) | 482 (93.2%) | 50 (98.0%) | 3.63 | 0.76, 65.2 | 0.208 | 4.10 | 0.85, 73.99 | 0.170 |
| **Animals** |  |  |  |  |  |  |  |  |  |
| Family contact with any animal | 484 (85.1%) | 439 (84.7%) | 45 (88.2%) | 1.35 | 0.60, 3.62 | 0.507 | 1.36 | 0.60, 3.67 | 0.494 |
| Animal faeces visible on the premises | 42 (7.4%) | 37 (7.1%) | 5 (9.8%) | 1.41 | 0.47, 3.48 | 0.490 | 1.44 | 0.47, 3.60 | 0.474 |
| Rodents sighted in household in past 7 days | 359 (63.1%) | 332 (64.1%) | 27 (52.9%) | 0.63 | 0.35, 1.13 | 0.118 | 0.58 | 0.32, 1.07 | 0.077 |
| **Food type^3^** |  |  |  |  |  |  |  |  |  |
| Cereal | 262 (46.3%) | 243 (47.1%) | 19 (38.0%) | — |  |  |  |  |  |
| Contains mixture of cereal, meat, fish vegetables or legumes | 148 (26.1%) | 125 (24.2%) | 23 (46.0%) | 2.35 | 1.24, 4.53 | 0.009 | 2.25 | 1.18, 4.35 | 0.015 |
| Contains dairy | 109 (19.3%) | 104 (20.2%) | 5 (10.0%) | 0.61 | 0.20, 1.58 | 0.346 | 0.61 | 0.20, 1.56 | 0.335 |
| Other | 47 (8.3%) | 44 (8.5%) | 3 (6.0%) | 0.87 | 0.20, 2.70 | 0.831 | 0.79 | 0.18, 2.48 | 0.720 |

^1^ N = 562

^2^ N = 568

^3^ N = 566

**Supplementary table 6.** Unadjusted and adjusted regression models for food-related risk factors and their association with Cryptosporidium detection in child food in Kenya.

| **Variable** | **Overall,**  **N = 569** | **No Cryptosporidium detection,**  **N = 539** | **Cryptosporidium detection,**  **N = 30** | **Unadjusted** | | | **Adjusted** | | |
| --- | --- | --- | --- | --- | --- | --- | --- | --- | --- |
|  |  |  |  | **uOR** | **95% CI** | **p-value** | **aOR** | **95% CI** | **p-value** |
| **Food purchasing** |  |  |  |  |  |  |  |  |  |
| Food vendor selected based on hygiene | 273 (48.0%) | 263 (48.8%) | 10 (33.3%) | 0.52 | 0.23, 1.12 | 0.104 | 0.56 | 0.24, 1.21 | 0.150 |
| Food vendor selected based on appearance | 99 (17.4%) | 95 (17.6%) | 4 (13.3%) | 0.72 | 0.21, 1.90 | 0.548 | 0.69 | 0.20, 1.83 | 0.496 |
| **Food storage and preparation** |  |  |  |  |  |  |  |  |  |
| Household owns a working refrigerator | 72 (12.7%) | 68 (12.6%) | 4 (13.3%) | 1.07 | 0.31, 2.84 | 0.908 | 1.62 | 0.43, 4.97 | 0.425 |
| Container used as preparation surface | 414 (72.8%) | 396 (73.5%) | 18 (60.0%) | 0.54 | 0.26, 1.18 | 0.111 | 0.57 | 0.26, 1.29 | 0.168 |
| **Cooking water** |  |  |  |  |  |  |  |  |  |
| JMP cooking water source - safely managed (Ref: Basic, limited, unimproved) | 76 (13.4%) | 73 (13.5%) | 3 (10.0%) | 0.71 | 0.17, 2.07 | 0.580 | 0.87 | 0.18, 3.12 | 0.844 |
| Point of use treatment of cooking water | 74 (13.0%) | 71 (13.2%) | 3 (10.0%) | 0.73 | 0.17, 2.14 | 0.616 | 0.82 | 0.19, 2.47 | 0.759 |
| **Sanitation** |  |  |  |  |  |  |  |  |  |
| JMP sanitation - at least basic (Ref: Limited and unimproved) | 115 (20.2%) | 111 (20.6%) | 4 (13.3%) | 0.59 | 0.17, 1.56 | 0.340 | 0.68 | 0.18, 2.15 | 0.544 |
| Human faeces visible on the premises | 14 (2.5%) | 13 (2.4%) | 1 (3.3%) | 1.40 | 0.08, 7.38 | 0.752 | 1.39 | 0.07, 7.82 | 0.757 |
| **Food preparation hygiene** |  |  |  |  |  |  |  |  |  |
| Reports handwashing before food preparation^1^ | 213 (37.9%) | 200 (37.6%) | 13 (43.3%) | 1.27 | 0.59, 2.66 | 0.529 | 1.40 | 0.65, 2.96 | 0.385 |
| Caregiver washes fruit and vegetables before use^2^ | 532 (93.7%) | 503 (93.5%) | 29 (96.7%) | 2.02 | 0.41, 36.5 | 0.496 | 2.37 | 0.47, 43.08 | 0.407 |
| **Animals** |  |  |  |  |  |  |  |  |  |
| Family contact with any animal | 484 (85.1%) | 455 (84.4%) | 29 (96.7%) | 5.35 | 1.12, 96.0 | 0.101 | 7.34 | 1.51, 132.30 | 0.053 |
| Animal faeces visible on the premises | 42 (7.4%) | 40 (7.4%) | 2 (6.7%) | 0.89 | 0.14, 3.12 | 0.878 | 1.16 | 0.18, 4.36 | 0.846 |
| Rodents sighted in household in past 7 days | 359 (63.1%) | 335 (62.2%) | 24 (80.0%) | 2.44 | 1.04, 6.67 | 0.056 | 2.64 | 1.10, 7.37 | 0.042 |
| **Food type^3^** |  |  |  |  |  |  |  |  |  |
| Cereal | 262 (46.3%) | 244 (45.5%) | 18 (60.0%) | — |  |  |  |  |  |
| Contains mixture of cereal, meat, fish vegetables or legumes | 148 (26.1%) | 137 (25.6%) | 11 (36.7%) | 1.09 | 0.49, 2.34 | 0.831 | 1.03 | 0.45, 2.24 | 0.939 |
| Contains dairy | 109 (19.3%) | 109 (20.3%) | 0 (0.0%) | NA |  |  | NA |  |  |
| Other | 47 (8.3%) | 46 (8.6%) | 1 (3.3%) | 0.29 | 0.02, 1.48 | 0.240 | 0.28 | 0.02, 1.46 | 0.230 |

^1^ N = 562

^2^ N = 568

^3^ N = 566

**Supplementary table 7.** Unadjusted and adjusted regression models for food-related risk factors and their association with *E. coli* composite detection in child food in Kenya.

| **Variable** | **Overall,**  **N = 569** | **No *E. coli* composite detection,**  **N = 539** | ***E. coli* composite detection,**  **N = 30** | **Unadjusted** | | | **Adjusted** | | |
| --- | --- | --- | --- | --- | --- | --- | --- | --- | --- |
|  |  |  |  | **uOR** | **95% CI** | **p-value** | **aOR** | **95% CI** | **p-value** |
| **Food purchasing** |  |  |  |  |  |  |  |  |  |
| Food vendor selected based on hygiene | 273 (48.0%) | 256 (47.5%) | 17 (56.7%) | 1.45 | 0.69, 3.09 | 0.330 | 1.35 | 0.64, 2.93 | 0.435 |
| Food vendor selected based on appearance | 99 (17.4%) | 96 (17.8%) | 3 (10.0%) | 0.51 | 0.12, 1.49 | 0.280 | 0.51 | 0.12, 1.49 | 0.276 |
| **Food storage and preparation** |  |  |  |  |  |  |  |  |  |
| Household owns a working refrigerator | 72 (12.7%) | 67 (12.4%) | 5 (16.7%) | 1.41 | 0.46, 3.52 | 0.499 | 1.04 | 0.31, 2.98 | 0.942 |
| Container used as preparation surface | 414 (72.8%) | 395 (73.3%) | 19 (63.3%) | 0.63 | 0.30, 1.40 | 0.237 | 0.64 | 0.30, 1.45 | 0.266 |
| **Cooking water** |  |  |  |  |  |  |  |  |  |
| JMP cooking water source - safely managed (Ref: Basic, limited, unimproved) | 76 (13.4%) | 73 (13.5%) | 3 (10.0%) | 0.71 | 0.17, 2.07 | 0.580 | 0.36 | 0.07, 1.29 | 0.151 |
| Point of use treatment of cooking water | 74 (13.0%) | 66 (12.2%) | 8 (26.7%) | 2.61 | 1.05, 5.88 | 0.027 | 2.40 | 0.95, 5.56 | 0.048 |
| **Sanitation** |  |  |  |  |  |  |  |  |  |
| JMP sanitation - at least basic (Ref: Limited and unimproved) | 115 (20.2%) | 108 (20.0%) | 7 (23.3%) | 1.21 | 0.47, 2.77 | 0.662 | 0.63 | 0.19, 1.88 | 0.427 |
| Human faeces visible on the premises | 14 (2.5%) | 14 (2.6%) | 0 (0.0%) | 0.00 |  | 0.989 | 0.00 | NA, 602885497976872370176.00 | 0.989 |
| **Food preparation hygiene** |  |  |  |  |  |  |  |  |  |
| Reports handwashing before food preparation^1^ | 213 (37.9%) | 202 (37.8%) | 11 (39.3%) | 1.06 | 0.47, 2.29 | 0.877 | 1.05 | 0.46, 2.27 | 0.908 |
| Caregiver washes fruit and vegetables before use^2^ | 532 (93.7%) | 503 (93.5%) | 29 (96.7%) | 2.02 | 0.41, 36.5 | 0.496 | 1.77 | 0.35, 32.35 | 0.583 |
| **Animals** |  |  |  |  |  |  |  |  |  |
| Family contact with any animal | 484 (85.1%) | 461 (85.5%) | 23 (76.7%) | 0.56 | 0.24, 1.44 | 0.191 | 0.31 | 0.11, 0.90 | 0.025 |
| Animal faeces visible on the premises | 42 (7.4%) | 39 (7.2%) | 3 (10.0%) | 1.42 | 0.33, 4.27 | 0.575 | 1.33 | 0.30, 4.18 | 0.657 |
| Rodents sighted in household in past 7 days | 359 (63.1%) | 346 (64.2%) | 13 (43.3%) | 0.43 | 0.20, 0.89 | 0.025 | 0.41 | 0.18, 0.88 | 0.023 |
| **Food type^3^** |  |  |  |  |  |  |  |  |  |
| Cereal | 262 (46.3%) | 252 (47.0%) | 10 (33.3%) | — |  |  |  |  |  |
| Contains mixture of cereal, meat, fish vegetables or legumes | 148 (26.1%) | 137 (25.6%) | 11 (36.7%) | 2.02 | 0.83, 4.97 | 0.117 | 1.96 | 0.80, 4.86 | 0.136 |
| Contains dairy | 109 (19.3%) | 100 (18.7%) | 9 (30.0%) | 2.27 | 0.88, 5.79 | 0.084 | 2.16 | 0.83, 5.56 | 0.108 |
| Other | 47 (8.3%) | 47 (8.8%) | 0 (0.0%) | NA |  |  | NA |  |  |

^1^ N = 562

^2^ N = 568

^3^ N = 566

**Supplementary table 8.** Summary of household characteristics by wealth index quintiles.

| **Asset-based characteristics included in the wealth index** | **Wealth Index Quintiles** | | | | | **Loading** |
| --- | --- | --- | --- | --- | --- | --- |
|  | **1 (N = 111)** | **2 (N =114)** | **3 (N =115)** | **4 (N =115)** | **5 (N =114)** |  |
| Household owns car | 0 (0%) | 0 (0%) | 0 (0%) | 0 (0%) | 37 (32%) | 0.734 |
| Household owns bicycle | 17 (15%) | 0 (0%) | 4 (3.5%) | 2 (1.7%) | 5 (4.4%) | -0.065 |
| Household owns motorbike | 2 (1.8%) | 0 (0%) | 17 (15%) | 8 (7.0%) | 22 (19%) | 0.010 |
| Household owns house | 0 (0%) | 0 (0%) | 51 (44%) | 15 (13%) | 54 (47%) | 0.481 |
| Improved wall material (Cement blocks or bricks) | 0 (0%) | 0 (0%) | 1 (0.9%) | 100 (87%) | 103 (90%) | 0.686 |
